# High-Throughput Adaptable SARS-CoV-2 Screening for Rapid Identification of Dominant and Emerging Regional Variants

**DOI:** 10.1101/2021.08.31.21262625

**Authors:** Zita Hubler, Xiao Song, Cameron Norris, Mehul Jani, David Alouani, Maureen Atchley, Lisa Stempak, Sarah Cherian, Christine Schmotzer, Navid Sadri

## Abstract

**Objectives:** Emerging SARS-CoV-2 variant strains can be associated with increased transmissibility, more severe disease, and reduced effectiveness of treatments. To improve the availability of regional variant surveillance, we describe a variant genotyping system that is rapid, accurate, adaptable, and able to detect new low-level variants built with existing hospital infrastructure.

**Methods:** We use a tiered high-throughput SARS-CoV-2 screening program to characterizes variants in a supra-regional health system over 76 days. Combining targeted qPCR and selective sequencing, we screen positive SARS-CoV-2 samples from all hospitals within our health care system for genotyping dominant and emerging variants.

**Results:** The median turnaround for genotyping was two days using the high-throughput qPCR-based screen, allowing us to rapidly characterize the emerging Delta variant. In our population, the Delta variant is associated with a lower CT value, lower age at infection, and increased vaccine breakthrough cases. Detection of low-level and potentially emerging variants highlights the utility of a tiered approach.

**Conclusions:** These findings underscore the need for fast, low-cost, high-throughput monitoring of regional viral sequences as the pandemic unfolds and the emergence of SARS-CoV-2 variants increases. Combing qPCR-based screening with selective sequencing allows for rapid genotyping of variants and dynamic system improvement.

**Key messages:**

- A tiered approach that uses qPCR-based screening to identify dominant variants and sequencing for unique variants maximizes throughput, turnaround time, and information gleaned from each sample.
- In our population, the Delta variant became dominant in less than a month and is associated with lower CT, lower age at infection, and increased breakthrough cases.
- We identified low-level variants, including the variant of interest B.1.621 and a Delta variant with an E484K mutation in our population using existing hospital infrastructure.

## INTRODUCTION

Over the last year, new variants of SARS-CoV-2 have emerged from all corners of the globe and some are associated with increased virulence, differing response to treatment, and the ability to evade vaccines. [1] New variants, combined with increased travel and variable vaccination status, can abruptly strain hospital resources.[2, 3] There is a need for quick identification and characterization of both dominant and low-level SARS-CoV-2 variants on a region-per-region basis to allow for appropriate allocation of resources in preparation for waxing and waning infection rates. Further, this identification system needs to be dynamic to monitor newly emerging variants that may become the dominant strain in a matter of weeks. While it would be ideal, use of sequencing for variant identification can be cost-prohibitive, low-throughput, and not broadly available in many clinical laboratories. By contrast, qPCR is available to most clinical laboratories and represents a speedy and cost-effective alternative to genotype common SARS-CoV-2 variants.[4–7] Several versions of qPCR-based variants screens detect common genetic alterations in the SARS-CoV-2 sequence to differentiate between variants. [4–7] Yet, qPCR-based screens are limited in their availability to detect newly emerging variants that are not identifiable with the existing genomic signatures. This work describes a tiered approach to SARS-CoV-2 variant screening that combines qPCR-based screening with the sequencing of select samples to monitor known variants and detect newly emerging variants quickly, accurately, and with flexible infrastructure investment.

## METHODS

### Samples

SARS-Cov-2 positive nasal or nasopharyngeal samples identified via routine clinical testing at one of 12 regional hospitals between May 21, 2021 and August 4, 2021 were included in the variant screening program. The program was developed to support regional and state variant surveillance efforts. The routine clinical testing platforms varied between regional hospitals and included: ID Now COVID-19 (Abbott), Simplexa COVID-19 Direct Kit (DiaSorin), Xpert Xpress SARS-CoV-2 (Cepheid), TaqPath COVID-19 Combo Kit (ThermoFisher Scientific), and Aptima SARS-CoV-2 (Hologic) Assays.

### RNA extraction and qPCR

All regional hospitals were instructed to send positive samples to a central lab performing the TaqPath COVID-19 Combo Kit assay to enable processing in a high-throughput 96 well plate format. RNA extraction and real-time qPCRs were performed per manufacturer’s instructions in a semi-automated fashion using a KingerFisher Flex System (ThermoFisher Scientific) and TaqPath COVID-19 Combo Kit (ThermoFisher Scientific) to assess for sufficient levels of ORF1ab, N-gene, and S-gene. The RNA plates were then transported to the sequencing laboratory to perform additional multiplexed qPCRs for regions of interest: ORF1del, N501Y, E484K, L452R. (For primers and more detailed methods, see Supplemental Table 1.) Data from all PCR plates were combined and integrated with patient demographic data. A deep learning model was used to identify any instances of errant cycle-threshold (Ct) calls. [4] Samples with N gene Ct greater than 33 were deemed to have an insufficient viral load for variant calling.

Vaccine breakthrough cases are defined as having occurred in patients who completed their vaccination series within the hospital system more than two weeks before the sample collection date.

### Variant preliminary calling

A preliminary probability matrix was calculated using publically available data that correlated the detection of specific viral genome markers with major WHO variants and CDC Variants of Concern (VoC)/Variants of Interest (VoI) using the primers described. An example of a probability matrix for publically available data is shown in Supplemental Table 2. The preliminary probability matrix was the basis for a ruleset that guides an automated preliminary calling algorithm. (Supplemental Table 3) The qPCR gene profile of each sample and the initial automated variant call are quickly reviewed and finalized. (Figure 1) Samples that have unclear expression profiles can be sequenced for further variant identification. (Figure 1)

**Figure 1:**
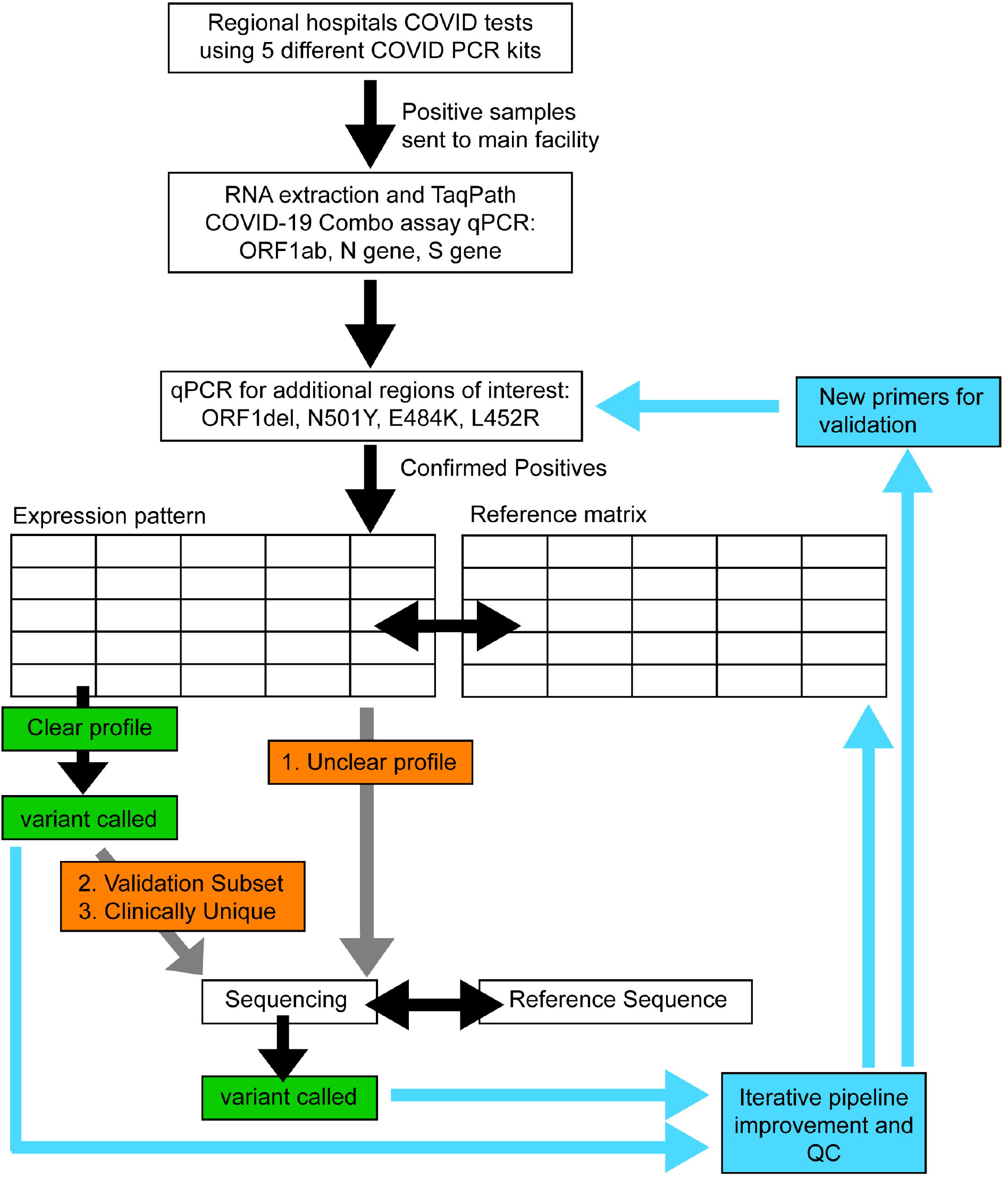
Schematic illustrating Dynamic Variant Calling Pipeline. Gray arrows indicate optional pathways, with examples of samples chosen for sequencing. Arrows and boxes coming from variant identification boxes show areas for iterative improvement and quality control of the pipeline. Confirmed positives are samples that have an N-gene Ct of less than 33.

### Sequencing

Samples were sequenced for one of three reasons: unclear expression profiles, variant sampling (pipeline validation), or clinically interesting cases (such as vaccine breakthrough). (Figure 1) The number of samples sequenced and sequencing timing is based on availability of clinical laboratory personnel and resources, and case numbers needed to comprise a sequencing batch. We performed whole-genome sequencing using the SARS-CoV-2 Research Panel (Ion Gene Studio S5 system, ThermoFisher Scientific) as previously described. [8] Pangolin (pangolin.cog-uk.io, v.3.1.3, 8/6/2021) and NextClad (clades.nextstrain.org, v.1.5.4) were used for lineage or clade assignment, respectively. Sequencing data was submitted to GeneBank (ncbi.nlm.nih.gov/genbank) for public use.

### Data Analysis

For this study, variant call from the qPCR-based Variant Screen and the sequencing results were combined and analyzed using Excel, R, and GraphPad Prism. Statistics were performed using GraphPad Prism. Data for the Delta+E484K samples were downloaded from GISAID (www.gisaid.org/) on 8/18/21. Data for the Cuyahoga County vaccination rates were downloaded from the Ohio Department of Health COVID-19 Dashboard (coronavirus.ohio.gov/wps/portal/gov/covid-19/dashboards) on 8/6/21.

## RESULTS

### Pipeline Performance

Between May 21, 2021 and August 4, 2021, 1150 positive COVID samples were processed in the qPCR-Based Screening Program, representing 2.0% of all COVID tests performed. (Figure 2) 99.5% of the screening results had an identifiable variant expression profile. Of all samples screened, 167 were subsequently sequenced for verification or clarification of variant calling. (Figure 2) Of note, based on our ruleset, the algorithm is not able to differentiate Pangolin lineages of the same variant, such as B.1.617.2 versus AY.3. However, it correctly classified the overall WHO variant as Delta. More granular identification of Pangolin lineages can be accomplished by incorporating novel primers and adjusting the ruleset (Figure 1). As part of our iterative improvement, we monitor the validity of the ruleset given the current variant distribution in our population. In the sequencing confirmed samples, for each variant called in the screen, the identification was correct 100% of the time (Figure 2). These results show that our qPCR screening is accurate and high-throughput, a necessary feature given the increasing rate of SARS-CoV-2 infections.

**Figure 2:**
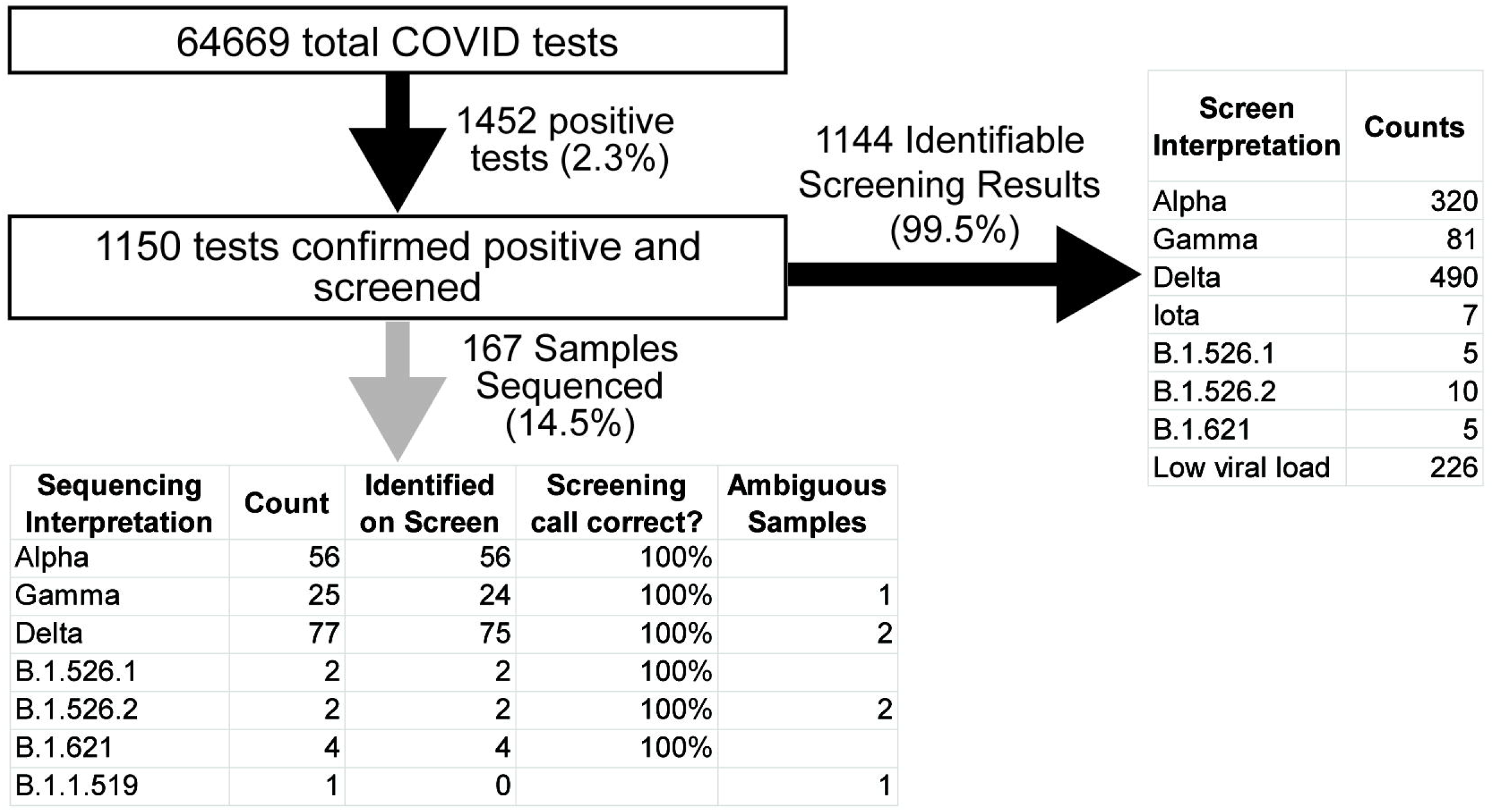
Flowchart of pipeline throughput over 76 days. Data collected from 5-21-21 to 08-04-21.

**Figure 3:**
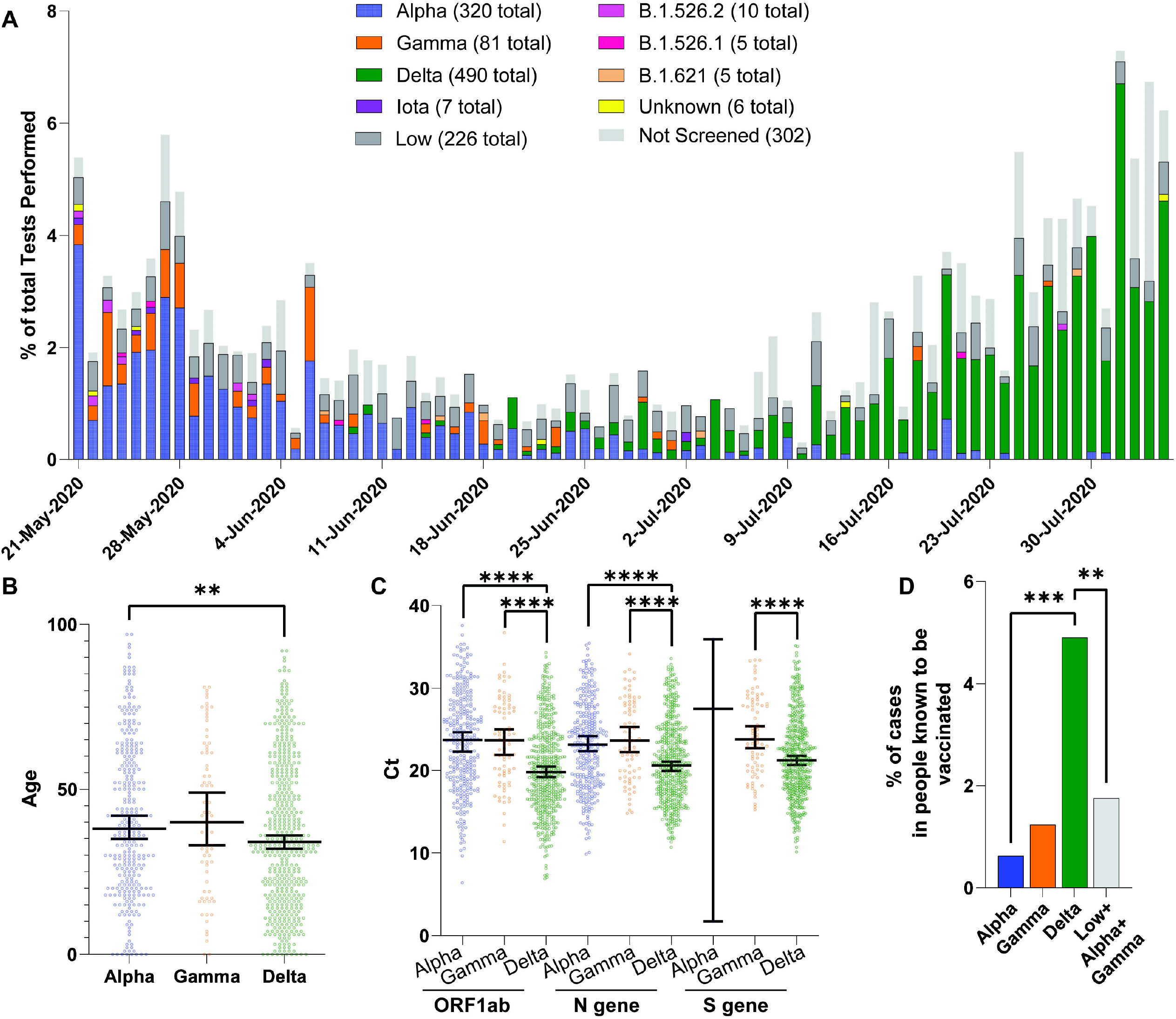
Tracking of Variants and Characterization of Delta. A) Percentage of each variant per total samples tested per day. Each bar corresponds to one day. Variants are color-coded and indicated in the legends. The totals indicated in the legend are the total count detected via the pipeline over 76 days. “Low” indicates samples with insufficient viral load for screening as measured by a high N gene Ct>33. “Not Screened” samples were not received at the main facility. B) Each dot indicates one sample, the y-axis is patient’s age, x-axis is the variant identified. Median and 95% confidence interval indicated with the bars. **, P<0.005, Mann Whitney two-tailed test. C) Variant versus Cycle threshold (Ct) for core markers: ORF1ab, N gene, and S gene. Each dot represents one sample. Median and 95% confidence interval indicated. ****, P<0.0001, One-way Anova with Tukey multiple comparison correction. D) Percentage of each variant that is in patients known to be vaccinated relative to all patients identified with that variant. ****, P < 0.0005, two-tailed Fisher exact test. **, P< 0.005, two-tailed Fisher exact test.

Another essential feature of SARS-CoV-2 variant surveillance is quick turnaround time. We found the median lag time before screening is two days after sample collection, with a degree of variability which may be attributable to differing transportation times from various facilities. (Supplemental Fig. 1A) By contrast, time to sequencing fluctuates, and may be delayed due to practical limitations such as personnel and equipment availability, as well as the number of available samples for a batch. Consequently, the median time between sample collection and sequencing was 14 days, substantially longer than the initial screen. (Supplemental Fig. 1A) This discrepancy in median screening versus sequencing time underscores the advantages of a rapid preliminary screen paired with subsequent sequencing.

### Variability in Variant Distribution over time

We analyzed the data for the variants called within the last 76 days, as this timeframe represents a relative plateau of vaccination initiation and completion in our county, reducing the temporal effects of changing vaccination rates in our community on the analyses performed. (Supplemental Fig. 1 B). We noted that in less than a month, the dominant strains shifted from being Alpha and Gamma to Delta. (Figure 2A) The transition rate between variants highlights the utility of a rapid variant calling algorithm. Overall, this data parallels the international trend; after the wave of Alpha swept our county, Delta arrived and was associated with an increase in cases that dominated all other strains in our region.[9, 10]

### Characterization of Delta Variant

Given the ongoing concern for Delta’s increasing viral load, infection of younger patients, and vaccine breakthrough cases, we wanted to characterize the impact of the Delta variant in our population.[9–13] We found a statistically significant decrease in the age of the patients testing positive for Delta compared to Alpha. (Figure 2B). Further, we found that Delta samples had cycle thresholds (Ct) significantly lower than Alpha and Gamma for the three SARS-CoV-2 markers assessed. (Figure 2C)[14] This may suggest an association between the Delta variant and increased viral load, as Ct has been used as an imperfect surrogate measure for viral load. Lastly, there was a statistically significant difference in the proportion of patients with each variant who were known to be vaccinated. [10] However, we noted a temporal association between the Alpha variant being dominant and the occurrence of low viral load vaccine-breakthrough events that were not genotyped. This suggested that difference in Ct values between Alpha and Delta could be confounding the analysis of vaccine-breakthrough. (Supplemental Fig. 1 C) Therefore, we assumed that all high Ct breakthrough cases were due to Alpha and Gamma, and despite this, Delta is associated with more vaccinated breakthrough cases. Together these results suggest that in our population, the Delta strain is infecting younger patients, with higher viral load, and more vaccine breakthrough cases.

### Identification of Novel Delta Variant

One advantage of our process is the ability to selectively use sequencing for samples that are most likely to be informative. While variants aside from Alpha, Gamma, and Delta, represented less than 3.6% of identified cases, these low-level variants can provide clues about the next more virulent or endemic strain. [2] (Figure 2A) Unbiased sample sequencing, especially when performed in limited numbers, may miss low-level variants. Our high-throughput qPCR screen identified some emerging low-level variants, such as 5 cases of B.1.621 (Figure 2A). However, novel or unique low-level variants require biased sequencing for further identification [2]. Our identification of a Delta variant with an E484K mutation exemplifies biased sequencing of samples with ambiguous qPCR screening results as a mechanism for identifying low-level variants. The sample had an unclear mutational signature on our qPCR screen that included an E484K and an L452R mutation. Within two days, we used sequencing and clustering to elucidate that the sample is a Delta variant based on its Pangolin lineage B.1.617.2 (ambiguity score: 0.9995) and Nextclade clade 21A. (Figure 4A) Sequencing confirmed the presence of an E484K mutation which is surprising because, this mutation is detected in less than 0.03% of Delta variants in GISAID (gisaid.org). Delta containing an E484K mutation is not prevalent in the USA but was detected in 56 Turkish samples, suggesting it may spread regionally and warrants additional surveillance. (Figure 4B) Interestingly, when phylogenetically arranged by Nextclade, our sample did not cluster with the samples from Turkey, making direct spread from Turkey less likely. (Supplemental Fig. 2). Instead was most similar to samples from the USA and Russia with, or without, an E484K mutation. (Supplemental Fig. 2) Overall, our detection of an uncommon Delta variant with an E484K mutation highlights the strength for monitoring low-level variants using a tiered qPCR and sequencing approach.

**Figure 4:**
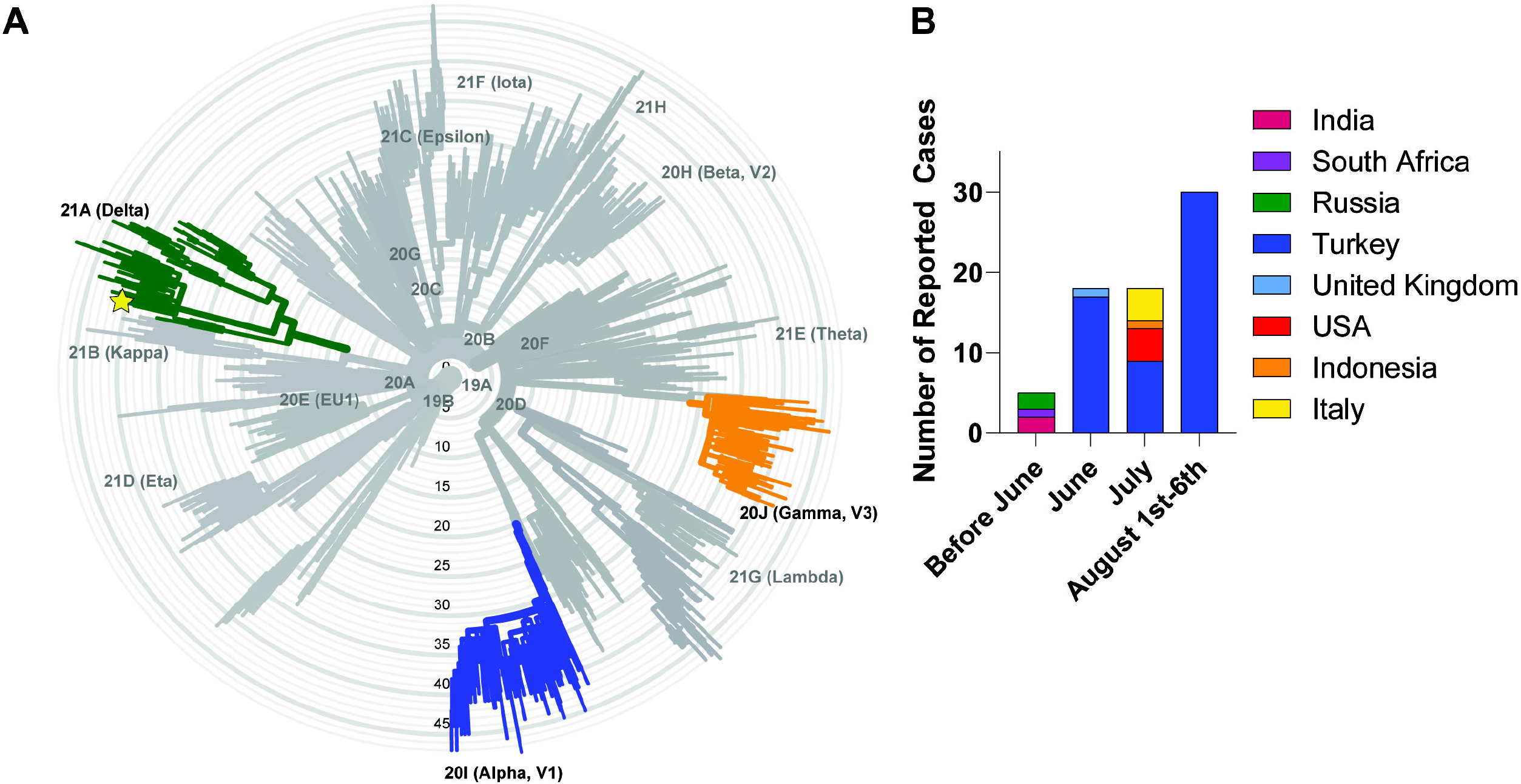
Identification of Delta + E484K. A) Phylogenetic tree showing the clade of the unknown sample (shown with the star). Radial axis is the number of mutations with Delta, Gamma, and Alpha highlighted. B) Number of reported cases of Delta +E484K in GISAID per country.

## DISCUSSION

As the COVID pandemic unfolds, the SARS-COV2 variants are evolving to be increasingly transmittable, have varying responses to treatment, and higher risk of evading existing vaccines. [1, 11] Information about newly emerging variants is vital for public health and hospital decision-making as they can overwhelm regional facilities and resources. [15] Therefore, healthcare systems need rapid and cost-effective variant detection to differentiate between subtly distinct and newly emerging variants. While no one molecular technique is ideal for all facilities, combining technologies in a tiered approach that combines qPCR screening and whole-genome sequencing allows for quick and accurate variant detection that is amenable to optimization.

qPCR-based testing is well-suited for identifying known variants with established mutational signatures but is limited in detecting novel variants. Clinical laboratories of all resource levels use qPCR-based variant screening due to ease of implementation and cost-efficiency. The widespread uptake of this technology is directly attributable to early work in the area that demonstrated the feasibility of variant identification using a combination of mutational markers, including all or some of the following: L452R, E484K, N501Y, H69_V70del. [5–7, 16, 17] Nevertheless, new variants with unique mutational signatures may not be identifiable with these existing qPCR-based screenings, demonstrating the need for further monitoring of low-level or novel variants.

Sequencing is the gold standard for identifying variants and the cornerstone for global monitoring of SARS-CoV-2 viral evolution. In fact, qPCR-based monitoring methods rely on global sequencing efforts to stay abreast with the newest viral strains. Resource-rich facilities can use sequencing for variant surveillance on a representative subset of unbiased samples. However, random sampling for sequencing may not be statistically geared to identify low-level variants on a regional basis. For example, identifying the Delta+E484K variant using unbiased sample selection for sequencing would necessitate sequencing thousands of un-mutated Delta variant samples. Turnaround time can be slower as sequencing runs may be delayed due to demands on laboratory resources when caseloads are high or until batches are filled when caseloads are low, limiting the utility of variant monitoring for real-time outbreak management. To enrich the sequencing data for information, some use targeted sequencing of mutational hotspots or restrict the population to hospitalized patients. [15, 18] Nevertheless, monitoring variants by exclusively sequencing is not feasible for most clinical laboratories due to resource limitations, preventing more widespread variant monitoring. Overall, both qPCR screening and unbiased population sequencing have some capacity to detect novel low-level variants on a regional basis. Using a tiered approach, we amplify the strengths of each, which may accelerate the identification of emerging clinically important strains.

We implemented a pipeline with a qPCR-based screen for the most common variants and subsequent sequencing of select samples. Examples of sequenced samples include: ambiguous qPCR expression profiles, clinically interesting cases, or quality control samples. Depending on available resources, sequencing can be performed in-house or commercially. In short, this tiered approach matches the strengths of each technology to the most informative samples. It is not intended to replace sequencing nor qPCR-based screening at all hospitals; it is an alternative approach for facilities to optimize their existing resources.

One advantage of the tiered approach is its capacity for dynamic feedback and iterative improvement. As the virus evolves, so too must the tests we use to detect it. In our workflow, the variant calling results can be used to adjust the primers and the screening ruleset to reflect variability in the regional level of variants. For example, if a variant of concern is noted in another country, such as Delta, we incorporate primers to detect it before the variant arrives in our population. Or, if a novel mutation is seen in the quality control samples from a dominant strain, the ruleset can be adjusted to account for the new subtype, such as for the Delta with an E484K. The current setup will not catch all mutations or new variants; instead, it is designed to be adaptable and broadly implementable. Given that the SARS-CoV2 genome is accumulating mutations and new variants are evolving, stagnant screening systems can become obsolete.[19–21] A tiered approach allows for continuous quality control and improvement to match the pace of variant evolution.

The merits of this system are exemplified by our timely detection of rare variants while maintaining high-throughput variant identification of the dominant strains. Due to the high-throughput nature of the qPCR screen, we identified the emerging Delta variant and, within the first few weeks, collected sufficient samples to perform preliminary characterization of it in our population. We found the Delta variant is associated with younger patient age, lower CT values, and increased vaccine breakthrough. As a testament to the utility of sequencing, we were able to detect a rare and emerging variant of Delta with an E484K mutation, which had not yet been described in our population and only a handful of times previously in the USA. Therefore, our approach demonstrates the strengths of both qPCR-based screening and sequencing and outlines the practical implementation of these tests using existing and strained clinical laboratory infrastructure during the middle of an evolving pandemic.

## Data Availability

Data sharing: All data relevant to the study are included in the article or uploaded as supplementary information. Sequencing data have been submitted to Genebank.

## Acknowledgements

We are grateful for the technical, medical, and administrative staff at University Hospitals Health System for their unwavering support and resilience of our SAR2- CoV-2 testing and variant identification programs.

