## Supplemental for "High-Throughput Adaptable SARS-CoV-2 Screening for Rapid Identification of Dominant and Emerging Regional Variants"

### SUPPLEMENTAL TABLES

**Supplemental Table 1:** Primers used for qPCR-based screen and qPCR conditions

| Plate | Primer Name | PCR Conditions | Primer Sequence | Citation/Source |
|---|---|---|---|---|
| First plate | ORF1ab | Per Manufacturer's instruction | | Taqpath COVID-19 Combo assay |
| | N gene | Per Manufacturer's instruction | | Taqpath COVID-19 Combo assay |
| | S gene | Per Manufacturer's instruction | | Taqpath COVID-19 Combo assay |
| Second Plate | ORF1del: Δ3675-3677 | 360 nM of each primer<br>80 nM of each probe<br>25°C, 2 min, 1X<br>52°C, 15 min, 1X<br>94°C, 2 min, 1X<br>94°C, 15 sec, 1X<br>57°C, 40 sec, 45X<br>68°C, 20 sec, 1X | Forward: TGCCTGCTAGTTGGGTGATG<br>Reverse:<br>TGCTGTCATAAGGATTAGTAACACT<br>Probe:<br>5CY5--GTTTGTCTG /TAO/<br>GTTTTAAGCTAAAAGACTGTG--3IAbRQSp | [6] |
| | S protein: N501Y | 360 nM of each primer<br>80 nM of each probe<br>25°C, 2 min, 1X<br>52°C, 15 min, 1X<br>94°C, 2 min, 1X<br>94°C, 15 sec, 1X<br>57°C, 40 sec, 45X<br>68°C, 20 sec, 1X | Forward: CTGAAATCTATCAGGCCGGTA<br>Reverse:<br>GAAAGTACTACTACTCTGTATGG<br>Probe:<br>56-FAM--TTTCCAACCCACTTATGGT--<br>3BHQ_1 | [7] |
| | S protein: E484K | 400 nM of each primer<br>50°C,10 min, 1X<br>95°C, 1 min, 1X<br>95°C, 2 min, 40X<br>55°C, 15 sec, 1X | Forward:<br>ACACCTTGTAATGGTGTTA<br>Reverse:<br>CTGGTGCATGTAGAAGTTCA<br>Probe:<br>SYBR Green | This manuscript |
| | S protein: L452R | 360 nM of each primer<br>80 nM of each probe<br>25°C, 2 min, 1X<br>52°C, 15 min, 1X<br>94°C, 2 min, 1X<br>94°C, 15 sec, 1X<br>57°C, 40 sec, 45X<br>68°C, 20 sec, 1X | Forward:<br>CTCTCTCAAAAGGTTTGAGATTAGACT<br>Reverse:<br>CTTGATTCTAAGGTTGGTGGTAA<br>Probe:<br>HEX--CCTAAACAATCTATACCGGTAATT--<br>3BHQ_1 | [7] |

**Supplemental Table 2:** Prevalence of genomic changes in GISAID samples 05/21/21-08/04/21

| Strain | Strain Type | # of GISAID sequences | ORF1del(%) | S:N501Y (%) | S: E484K (%) | S: L452R (%) |
|---|---|---|---|---|---|---|
| Alpha (B.1.1.7) | VoC | 115403 | 96.5 | 97.8 | 0.8 | 0.1 |
| Beta (B.1.351) | VoC | 2988 | 90.9 | 87.6 | 87.2 | 0.3 |
| Gamma (P.1) | VoC | 18685 | 97.6 | 95.3 | 94.9 | 0.0 |
| Delta (B.1.617.2, AY.3) | VoC | 418169 | 0.1 | 0.1 | 0.0 | 98.0 |
| Eta (B.1.525) | VoI | 560 | 95.4 | 0.4 | 97.3 | 0.0 |
| Ito (B.1.526) | VoI | 1949 | 96.9 | 0.1 | 78.0 | 0.0 |

**Supplemental Table 3:** Example ruleset for calling of Variants on qPCR-based Screen

| ORF1ab | N gene | S gene | ORF1 del: Δ3675-3677 | S: N501Y | S: E484K | S: L452 | Interpretation |
|---|---|---|---|---|---|---|---|
| CT< 40 | CT<40 | (S gene CT) – (N gene CT) =< 6 | CT<40 | CT<40 | (E484K CT) – (N gene CT) =<5 | CT<40 | |
| TRUE | TRUE | FALSE | FALSE | TRUE | FALSE | FALSE | **Alpha (B.1.1.7)** |
| TRUE | TRUE | FALSE | FALSE | TRUE | TRUE | FALSE | **Alpha (B.1.1.7+E484K)** |
| TRUE | TRUE | TRUE | FALSE | TRUE | TRUE | FALSE | **Gamma or Beta** |
| TRUE | TRUE | TRUE | TRUE | FALSE | FALSE | TRUE | **Delta** |
| TRUE | TRUE | FALSE | FALSE | FALSE | TRUE | FALSE | **Eta** |
| TRUE | TRUE | TRUE | FALSE | FALSE | TRUE | FALSE | **Iota** |
| TRUE | TRUE | TRUE | FALSE | FALSE | FALSE | TRUE | **B.1.526.1** |
| TRUE | TRUE | TRUE | FALSE | FALSE | FALSE | FALSE | **B.1.526.2** |
| TRUE | TRUE | TRUE | TRUE | TRUE | TRUE | FALSE | **B.1.621** |
| TRUE | TRUE | TRUE | TRUE | FALSE | TRUE | FALSE | **P.2** |

### SUPPLEMENTAL FIGURES

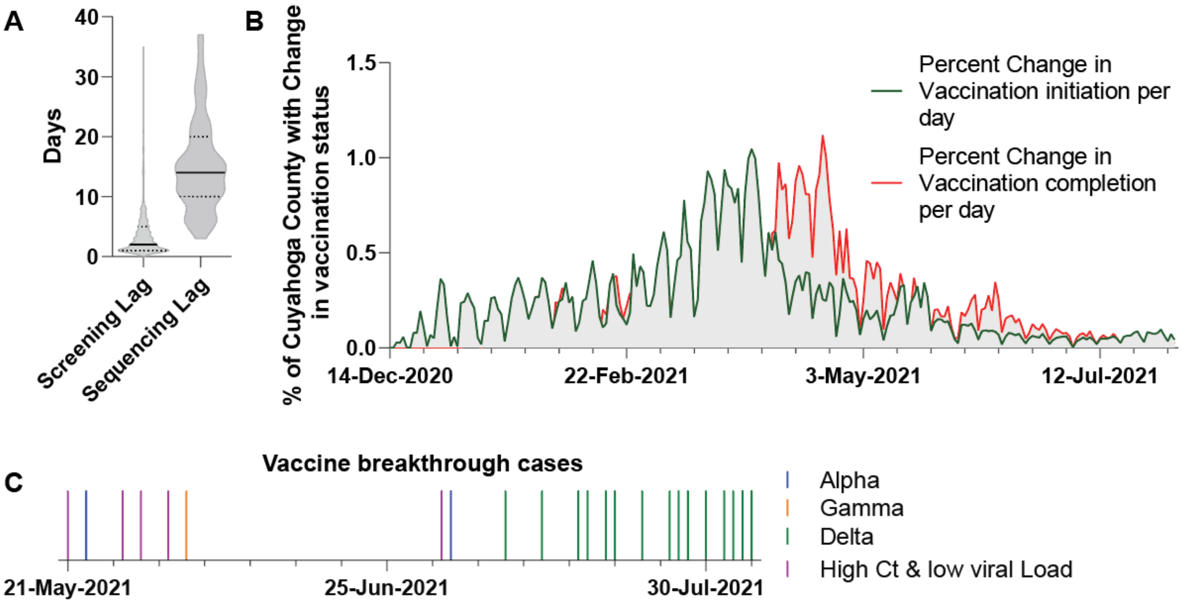


**Supplemental Figure 1:** A) Elapsed time between collection, screening and sequencing for all samples analyzed. Solid line indicates the median, quartiles indicated with dotted lines. B) Cuyahoga County vaccination initiation and completion rates as a percentage of the population. C) Incidence of positive SARS-CoV-2 tests over time per variant. Each line indicates one positive result. Colors indicate the variant identified or high Ct, respectively.

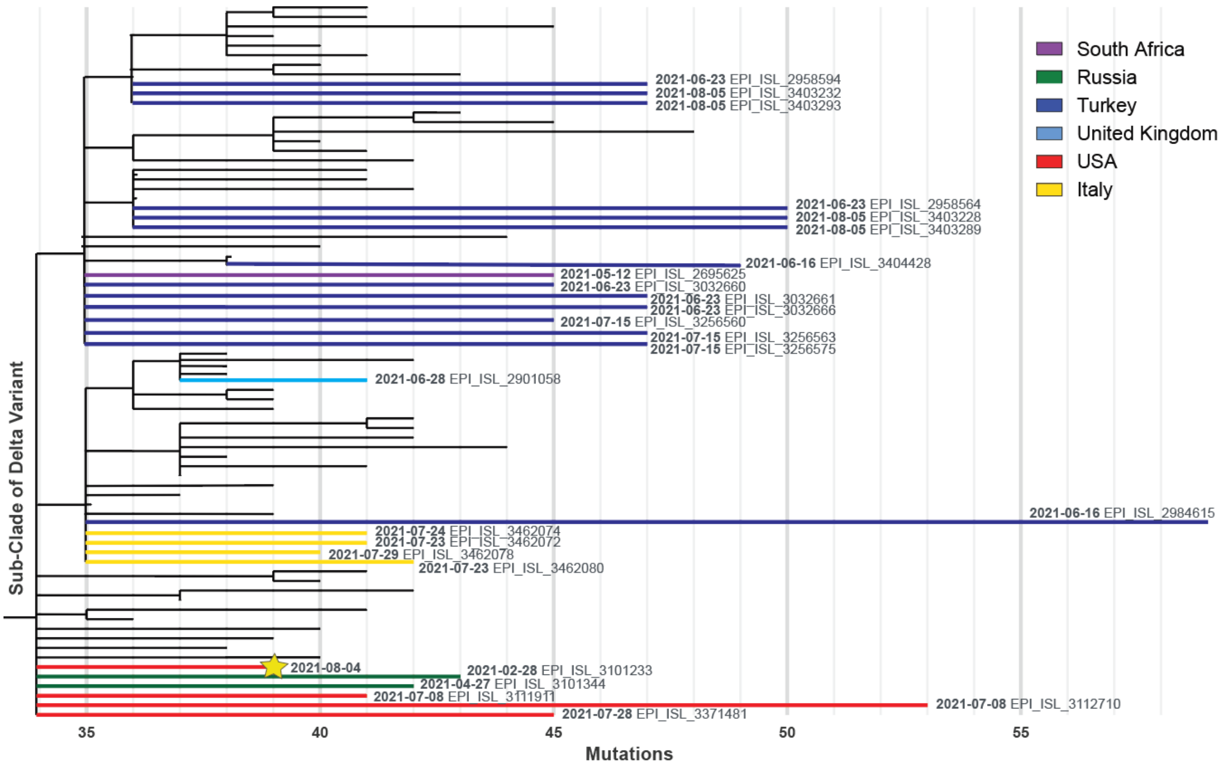


**Supplemental Figure 2:** Subset of Nextclade phylogenetic tree containing the Delta+E484K sample (indicated with star) and most closely related Delta+E484K samples. Delta variant reference samples are shown in black lines. For Delta+E484K samples, country of origin labeled with colors per legend, date sample was collected in bold, and Accession ID's indicated.

### REFERENCES (as per main text)


6. Vogels, C.B.F., et al., Multiplex qPCR discriminates variants of concern to enhance global surveillance of SARS-CoV-2. PLoS Biol, 2021. 19(5): p. e3001236.

7. Wang, H., et al., Multiplex SARS-CoV-2 Genotyping Reverse Transcriptase PCR for Population-Level Variant Screening and Epidemiologic Surveillance. J Clin Microbiol, 2021. 59(8): p. e0085921.
